# Can far become near? The effect of approaching visual stimuli on tactile processing in fibromyalgia patients and controls

**DOI:** 10.64898/2026.11.18.26368406

**Authors:** Annick L. De Paepe, Geert Crombez, Valéry Legrain

## Abstract

Within the space closely surrounding us (i.e. the peripersonal space), stimuli on the body surface are integrated with stimuli in the external world. This enables us to interact with the world, and to defend ourselves against potentially threatening objects approaching our body. It has been shown that attention towards stimuli approaching us can differ depending on, for example, the level of anxiety. The present study investigated whether fibromyalgia (FM) patients have a heightened attention to stimuli entering the peripersonal space compared to healthy control participants. This was done by investigating the differential influence of dynamical visual stimuli approaching the body on tactile processing for control participants versus FM patients. For control participants we found, in accordance with previous research, that visual stimuli presented near as opposed to far from the body influenced tactile processing more. For FM patients this difference was less clear, possibly indicating that FM patients have more attention for potentially threatening stimuli at further distance. The curves describing the reaction times along the continuous spatial range (from near to far) indicated that FM patients have a heightened attention for stimuli entering the peripersonal space compared to controls. However, as this difference was only found when curve-fitting the data, we argue that our results should be interpreted with caution and need further corroboration and replication.

## 1. Introduction

The peripersonal space is the space immediately surrounding our body in which we can interact with the external world (Graziano & Cooke, 2006; Rizzolatti, Fadiga, Fogassi, & Gallese, 1997). This interaction is only possible when stimuli on the body space are integrated with stimuli occurring in the external world. Indeed, it has been shown that external stimuli (e.g., visual stimuli) occurring within the peripersonal space, are integrated with somatosensory stimuli on the body space, whereas stimuli occurring beyond the peripersonal space (i.e. in extrapersonal space) are not (or to a lesser extent) (for nociceptive stimuli: De Paepe, Crombez, & Legrain, 2015; De Paepe, Crombez, Spence, & Legrain, 2014; for tactile stimuli: see Spence & Driver, 2004, for a review).

Besides the possibility to interact with the world, this ability also provides us with a protective mechanism, allowing us to, for example, brush away an insect before it can sting us. In this sense, the peripersonal space can be considered a ‘safety margin’ around our body, which we are scanning for potentially threatening objects approaching our body and which allows for the swift preparation of defensive reactions against intruders (Legrain & Torta, 2015; Legrain, 2011; Moseley, Gallace, & Spence, 2012). Evidence in favor of this view has been provided by cortical stimulation studies. When the brain areas associated with multisensory processing in monkeys are electrically stimulated, defensive arm movements and withdrawing of the arm and the head are observed (Graziano & Cooke, 2006). In humans, it has been shown that three-dimensional visual distractors rapidly approaching participants’ hands modulated corticospinal excitability over the primary motor cortex (Makin, Holmes, Brozzoli, Rossetti, & Farnè, 2009).

It has been argued that the boundaries of the peripersonal space are not fixed. This has been demonstrated by the fact that the peripersonal space can be ‘extended’ through tool-use (Bassolino, Serino, Ubaldi, & Làdavas, 2010; Farnè, Serino, & Làdavas, 2007; Maravita & Iriki, 2004; Serino, Bassolino, Farnè, & Làdavas, 2007). For example, Farnè & Làdavas (2000) used a crossmodal paradigm, used to reveal visuo-tactile interaction, in patients with tactile extinction. These patients can detect tactile stimulation to their left hand in isolation, but they fail to report the left-hand stimulation when the right hand is concurrently stimulated (unimodal extinction) or when a right visual stimulus is presented near the right hand (crossmodal extinction). Farnè & Làdavas (2000) assessed crossmodal visual-tactile extinction by presenting visual stimuli far from the patients’ ipsilesional hand (∼30 cm), near the distal edge of a rake that was held in their hand. The study revealed that following the use of the rake to retrieve distant objects, crossmodal extinction was more severe as opposed to when the rake was not used. This effect only lasted for a few minutes after tool-use. This shows that the use of a tool can increase the spatial extent of the representation of the peri-hand space to incorporate a tool. Other studies showed that the peripersonal space can be modulated by social interactions with others (Teneggi, Canzoneri, di Pellegrino, & Serino, 2013), or by anxiety (Taffou & Viaud-Delmon, 2014). Taffou & Viaud-Delmon (2014) investigated the extent of the peripersonal space in the presence of threatening (dog growling) and non-threatening (sheep bleating) auditory stimuli looming from the rear hemifield in non-fearful and dog-fearful individuals. They found that the peripersonal space of dog-fearful individuals was enlarged when a threatening sound as opposed to a non-threatening sound approached them. This effect was not found in non-fearful individuals. The authors argued that this enlargement of the peripersonal space in the presence of feared elements would be adaptive as it provides more time to prepare for a defensive response, and as such it fits with the proposed protective function of the peripersonal space.

In the present study, we investigated whether attention to stimuli approaching the body can be different for fibromyalgia patients compared to healthy control participants. Fibromyalgia is characterized by chronic widespread pain and tenderness in muscles and joints. Studies have indicated that FM patients demonstrate an exaggerated response to experimentally induced noxious stimuli, compared to other groups (Granges, Gibson, Littlejohn, & Helme, 1993; Granges & Littlejohn, 1993; Lautenschlager, Seglias, Bruckle, & Muller, 1991; Scudds, Rollman, Harth, & McCain, 1987; Tunks, Crook, Norman, & Kalaher, 1988). Interestingly, some studies suggest that patients with chronic pain not only have an over-responsiveness to painful stimuli, but also to stimuli in other modalities (e.g., sound) (Crombez, Van Damme, & Eccleston, 2005; McDermid, Rollman, & McCain, 1996). For example, Hollins et al. (2009) exposed FM patients, patients with temporomandibular disorders (TMD) and control participants to a set of pressure stimuli and a set of auditory stimuli and asked to rate the intensity, as well as the unpleasantness of each stimulus. FM patients rated the intensity and the unpleasantness of the stimuli significantly higher than the two other groups in both modalities, although the effect was stronger for the cutaneous stimuli. The origin of FM is still unknown, but the pattern of complaints and perceptual amplification associated with FM suggests dysfunctions in central processes (Wall, 1993). One hypothesis is that patients with FM are characterized by ‘hypervigilance’, referring to ‘a habit to attend to somatic distress signals’ (Chapman, 1978). It is proposed that this results from a cognitive process in which FM patients are concerned about, and therefore closely monitor, those sensations that could accompany or warn impending pain, leading to an increase of response to all stimuli of that type (Hollins et al., 2009). It has to be noted that hypervigilance is only one mechanism that may account for the research findings demonstrating hypersensitivity in FM patients, such as the study of Hollins et al. (2009). Other processes such as central sensitization (e.g., Arendt-Nielsen & Henriksson, 2007; Staud, Robinson, & Price, 2007) have also been suggested to account for lowered pain threshold and tolerance levels in FM patients. To talk about ‘hypervigilance’ one should demonstrate the involvement of attentional processes, like vigilance and scanning towards those sensations that could accompany or warn impending pain (Crombez et al., 2005; Damme et al., 2009).

Here we want to investigate whether FM patients have a heightened attention for stimuli entering the peripersonal space or whether they scan a larger share of the external space for potentially salient and threatening information. To investigate this hypothesis, we studied the influence of dynamical visual stimuli on tactile processing in healthy control participants and FM patients. A visual stimulus was either approaching or receding the participant’s left or right hand. At different temporal delays from the onset of the visual stimulus, a tactile stimulus was applied either at the same or the opposite hand, so that it was presented when the visual stimulus was perceived at varying distances from the hand. Participants were asked to respond as fast as possible at which side they perceived a tactile stimulus. We expected that RTs to tactile stimuli would progressively decrease as a function of the perceived approach of the visual stimulus. Conversely, we expected RTs to increase as a function of the perceived recession of the visual stimulus. This effect should be more pronounced when the light was presented at the same as opposed to the opposite side of the tactile stimulation. The best fitting curves of the RTs as a function of the perceived position of the visual stimuli in space were studied in order to compare the influence of approaching visual stimuli on tactile processing between FM patients and healthy controls.

## 2. Methods

### 2.1. Participants

Forty patients with FM and a control group of forty-one participants matched for age, sex and level of education were recruited. FM patients were recruited via the Multidisciplinary Pain Clinic of Ghent University Hospital. Inclusion criteria were a diagnosis of FM (Wolfe et al., 2010), an age between 18 and 65 years, normal or corrected-to-normal eyesight (e.g., lenses, glasses), normal or corrected-to-normal hearing (e.g., hearing aid), and Dutch speaking. Exclusion criteria were suffering from neurological problems (e.g., epilepsy) and insensitivity on the hands. Potential participants were informed about the possibility of participating by means of a poster in the waiting room, information given by the physician and information letters. When they agreed to participate, they received a phone call from the researcher providing details about the study. 2 patients were excluded from the sample, because they performed poorly on the task (see section 3.3.). The remaining 38 patients reported pain complaints for, on average, 15 years (SD = 12 years). Pain was reported on an average of 171 days (SD = 26) over the last 6 months. 63% of them were not working because of the pain and received a monthly allowance. On average, the FM group reported being unable to perform daily activities (work, household) on 87 days (SD = 72) over the last 6 months.

The control participants were recruited via advertisement in local newspapers (e.g., Zone09), social media (Twitter, Facebook) and flyers distributed around the university campus, the local library, etc. Inclusion criteria were an age between 18 and 65 years, normal or corrected-to-normal eyesight (e.g., lenses, glasses), normal or corrected-to-normal hearing (e.g., hearing aid), and Dutch speaking. Exclusion criteria were suffering from pain of a severe intensity, meeting the criteria of FM according to the questionnaire of Wolfe et al. (2010), suffering from neurological problems and insensitivity on the hands. Pain of a severe intensity was operationalized as a grade score on the Von Korff scale (Von Korff, Ormel, Keefe, & Dworkin, 1992) greater than or equal to three, indicating the experience of pain that is at least moderately disabling. Three control participants were excluded because of this exclusion criterion. One other participant was excluded because he/she fulfilled the criteria for FM as described in Wolfe et al. (2010).

The final sample consisted of 38 patients and 37 control participants. Demographics and scores on the Von Korff scale (Von Korff et al., 1992) and the FM questionnaire (Wolfe et al., 2010) for both groups can be found in Table 1.

**Table 1.** Demographics and pain scores for 38 patients and 37 control participants. \**p* < 0.05, \*\*\**p* < 0.001.

|  | **Patients<br>(N = 38)** | **Controls<br>(N = 37)** | **t/χ<sup>2</sup>** | ***P*** |
|---|---|---|---|---|
| **Age** | 44.76 (9.14) | 45.38 (10.25) | -0.27 | .78 |
| **Sex (% female)** | 92% | 89% | 0.19 | .70 |
| **Education level** |  |  | 5.34 | 0.25 |
| Primary school (< 12 years) | 3% | 0% |  |  |
| Lower secondary school (< 15 years) | 21% | 5% |  |  |
| Higher secondary school (< 18 years) | 39% | 54% |  |  |
| High school | 34% | 38% |  |  |
| University | 3% | 3% |  |  |
| **Handedness (% right handed)** | 87% | 92% | 0.50 | .70 |
| **% unmarried/divorced/widow** | 39% | 54% | 1.07 | .30 |
| **% having children** | 79% | 59% | 2.49 | .11 |
| **Unemployment** | 74% | 24% | 16.35 | <.001<sup>***</sup> |
| **% participants reporting poor state of health** | 15% | 0% | 6.35 | .02<sup>*</sup> |
| **FM questionnaire (Wolfe et al., 2010):** |  |  |  |  |
| ***Widespread Pain Index (WPI)*** | 10.45 (4.18) | 1.38 (1.46) | 12.62 | <.001<sup>***</sup> |
| ***Severity Symptom (SS) scale*** | 7.61 (2.19) | 2.89 (1.81) | 10.18 | <.001<sup>***</sup> |
| **Grade score<br>(Von Korff et al., 1992)** | 3.24 (1.13) | 0.11 (0.31) | 15.59 | <.001<sup>***</sup> |

The experiment lasted approximately 1.5 hours and was part of a larger protocol that had been approved by the Ethical Committee of the Ghent University Hospital (see http://hdl.handle.net/1854/LU-7032736). At the end of the experiment, the participants received 25 euros as reimbursement for their expenses.

### 2.2. Stimuli and apparatus

Vibrotactile stimuli were presented by means of two resonant-type actuators (C-2 TACTOR, Engineering Acoustics, Inc., Florida, http://www.eaiinfo.com), consisting of moving magnet linear actuators in a housing of 3.05 cm diameter and 0.79 cm high, with a skin contactor of 0.76 cm diameter. The tactile stimuli had a frequency of 300 Hz, and a duration of 10 ms. The intensity of the tactile stimuli was determined for each participant individually by means of a random staircase procedure. Twenty tactile stimuli were presented to one of the hands (i.e. the ‘reference hand’, for half of the participants this was the left hand, for the other half the right hand) (the intensity started between 0 and 0.27 Watt) and self-reports were collected on an 11-point Likert scale (0 = ‘not intense at all’ to 10 = ‘very intense’). The intensity that elicited an average rating of 7 was selected as the intensity of the tactile stimulation for the reference hand during the experiment. The perceived stimulus intensity of the other hand was matched (Weinstein, 1968). This was done by means of a double random staircase procedure, based on the ‘simple up-down method’ of Levitt (1971). Twenty-four stimuli on this hand were judged relative to the stimulus on the reference hand on a 5-point Likert scale (1 = ‘more than less strong’, 2= ‘less strong’, 3= ‘equally strong’, 4= ‘stronger’, 5= ‘much stronger’). The intensity that elicited an averaged rating of 3 was used as the intensity of the stimulus on the other hand.

The visual stimuli were presented by means of fourteen green light-emitting diodes (LEDs). One red LED was used for fixation.

The participants sat on a chair in a dimly illuminated, sound-attenuated room, with their head position fixed in a chin rest. The height of the chin rest was individually adapted. Participants rested their arms on the table in front of them, and placed their hands, palm downward on the table. The distance between the participants’ hands and their trunk, as well as the distance between the participants’ index fingers was 40 cm. In total 14 LEDs were positioned at different distances from the hands. Seven LEDs were positioned in the left side of space, and 7 LEDs in the right side of space. At both sides, the first LED was positioned in between thumb and index finger, the next six LEDs were positioned on a straight line, one in front of the other with 12 cm in between successive LEDs, so that the last LED was 72 cm in front of the first LED. On each trial, the LEDs on one side were successively illuminated, creating the illusion of a light coming closer towards the participant (the last LED, at 72 cm distance, was first illuminated), or going further away from the participant (the first LED, in between thumb and index finger, was first illuminated). Each LED was illuminated for 280 ms, and each offset was immediately followed by the illumination of the next LED, so that the total dynamical visual stimulus had a duration of 1960 ms. A red fixation LED was positioned in between the LEDs in left and right space, 36 cm in front of the first LEDs. This fixation LED was illuminated at the beginning of each trial and was turned off for 1s after a response was given.

### 2.3. Procedure

The experiment started by illuminating the LEDs one by one. Participants were asked to look at the fixation LED and to indicate verbally at which side of space a light was illuminated (i.e. “left” or “right”). This was done to ensure that participants could see all the LEDs. Next, participants completed a practice phase of 14 trials, in which they had to achieve 90% correct performance in order to proceed with the experiment. All participants reached this criterion.

The procedure is based on a study of Canzoneri, Magosso, & Serino (2012). Each trial started with the illumination of the fixation LED for 1s. Thereafter the dynamical visual stimulus started. At different temporal delays after the onset of the first visual stimulus, a tactile stimulus could be presented: T1, a tactile stimulus was administered 170 ms from first light onset; T2, 450 ms from first light onset; T3, 730 ms from first light onset; T4, 1010 ms from first light onset; T5, 1290 ms from first light onset; T6, 1570 ms from first light onset; T7, 1850 ms from first light onset. This was true both for the approaching and the receding light. In this way, the light was perceived at different locations with respect to the body at the moment the tactile stimulation occurred. For example, when the light was approaching it appeared close at high temporal delays. Conversely, when the light was receding, it appeared close at low temporal delays (see Figure 1).

**Figure 1.**
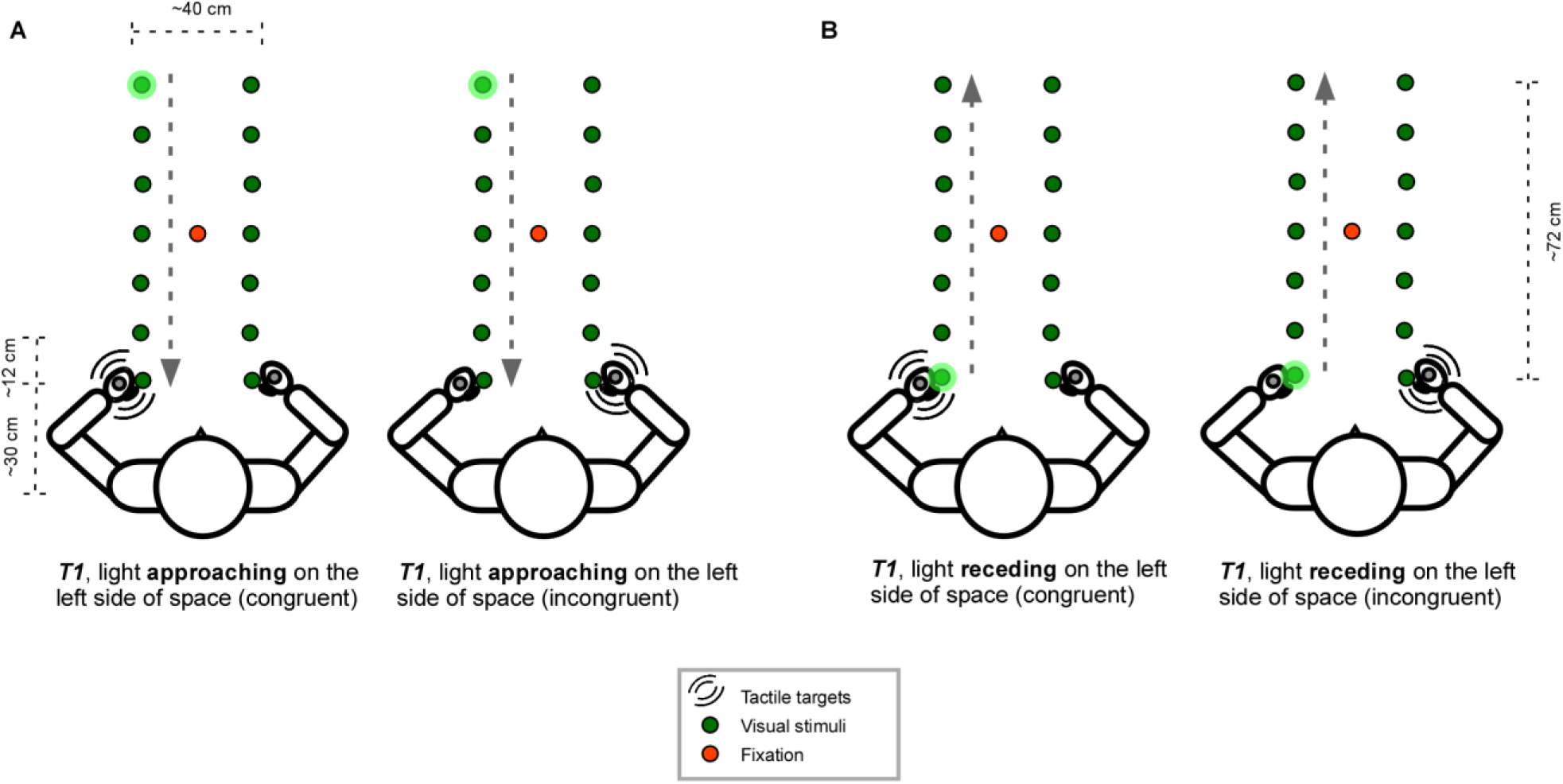
Experimental set-up. **A.** A light is *approaching* the participant at the left side of space. At T1 (170 ms from light onset) the participant gets a tactile stimulus applied to the left hand (left figure, congruent to the side of space where the light is presented) or to the right hand (right figure, incongruent to the side of space where the light is presented). At that time, the light is at 72 cm from the participant’s hand. **B.** A light is *receding* from the participant’s hand, so that the light is now in between the thumb and the index finger at the time of stimulation. At T1 (170 ms from light onset) the participant gets a tactile stimulus applied to the left hand (left figure, congruent to the side of space where the light is presented) or to the right hand (right figure, incongruent to the side of space where the light is presented).

The experiment consisted of 8 blocks of 56 trials each. The trials were created by crossing the moving direction of visual stimulus (approaching vs. receding) with the congruency of the visual and tactile stimulus (congruent vs. incongruent), the side at which the visual stimulus was presented (left/right side of space) and the 7 different temporal delays (T1 - T7). 1/8 of the trials (i.e. 7 trials) per block were randomly assigned as catch trials, in which no tactile stimulus was presented.

Participants were instructed to keep their gaze on the fixation LED during the whole block. They were asked to respond as fast and accurately as possible. Responses were given by means of two foot pedals, one positioned beneath the toes, and one beneath their heel. Participants were instructed to keep the foot pedals depressed during the experiment, and to lift either their toes or their heel to respond. Half of the participants responded with their left foot, the other half with their right foot. The response mapping (toe = left hand, heel = right hand, or vice versa) was counterbalanced between participants. Participants were informed that the visual stimulus was unpredictive for the position of the subsequent tactile target. To mask any noise produced by either the foot pedals or the tactile stimuli, participants wore headphones (Sennheiser, HD201). The experiment took on average 75 minutes to complete.

### 2.4. Measures

Because participants were very accurate in performing the task (see section 3.3.), performance was only analyzed in terms of the reaction time (RT). Only RTs from correct trials were considered in the analyses. Inspection of quantile-quantile plots suggested that the data was not normally distributed and that an Inverse-Gaussian transformation was optimal both for the data of control participants and of FM patients (Ratcliff, 1993). After transforming the data, RTs exceeding three times the median absolute deviation (MAD) (Leys, Ley, Klein, Bernard, & Licata, 2013) were considered outliers and were trimmed from the analyses separately for FM patients and control data (respectively 0.8 and 1.4% of trials on average over all conditions). The inversely transformed RTs were multiplied by −1000 so that coefficients would have the same sign as for models with untransformed RTs, at the same time avoiding very small values and a too restricted range for the dependent variable. Mean RTs were calculated for every temporal delay, for congruent and incongruent trials, and for approaching and receding visual stimuli, creating 28 different conditions for each group (control vs. patient).

Between each block participants were asked to rate the intensity of the stimulation for the left and the right hand on a 10-point numerical rating scale (going from 0 = felt nothing, over 5 = fairly intense, to 9 = very intense). The equivalence of the average intensity for the left compared to the right hand for both groups was assessed using paired samples t-tests. Moreover, the equivalence of the self-reported intensity ratings was compared between both groups using one-sample t-tests.

After the experiment participants were asked to indicate how threatening they thought the visual lights were both when the light was approaching, and when the light was receding, on a scale from 0 (not at all) to 10 (extremely). The perceived threat score was compared for approaching and receding visual stimuli for both groups using Wilcoxon signed rank test for paired samples. Moreover, the perceived threat scores were compared for both groups using Wilcoxon signed rank test for unpaired samples.

### 2.5. Analyses

Based on previous research we expected the lights approaching the stimulated hand to have a stronger spatially dependent effect on tactile processing than either receding lights or lights approaching the non-stimulated hand (Canzoneri et al., 2012; De Paepe, Crombez, & Legrain, under review; Teneggi et al., 2013). If this is true, it would be especially interesting to compare control and patient data for the trials in which the lights were approaching the stimulated hand. To check this, we first analyzed control and patient data separately and checked whether approaching and receding lights differentially affected reaction times for both groups (Step 1: effect of visual stimulus direction). Next, to increase power and for the sake of parsimony, we only compared control and patient data for trials in which the lights were approaching participants’ stimulated hand (Step 2: effect of group).

Results were analyzed using linear mixed effects models as implemented in the package “Linear and Nonlinear Mixed Effects Models” (Pinheiro & Bates, 2000). Linear mixed effects models account for the correlations in within-subject data by estimating subject-specific deviations (or random effects) from each population-level factor (or fixed factor) of interest (see West, Welch, & Galecki, 2007, for an elaboration). The outcome variable of interest was the RT. During Step 1 the independent variables were the visual stimulus direction (approaching versus receding lights), the congruency (congruent vs. incongruent), and the temporal delay (T1 to T7). These were manipulated within subjects. During Step 2 the independent variables were group (control vs. patient) and temporal delay (T1 to T7), which were respectively between - and within-subjects variables. Each analysis required three steps. First, all relevant factors and interactions were entered in the model as fixed factors, and we assessed whether it was necessary to add a random effect for each of the fixed (within subject) factors in the analysis: If a random effect significantly increased the fit of the model, it was included in the final model. By default, a random effect was added introducing adjustments to the intercept conditional on the Subject variable. In the second step, we searched for the most parsimonious model that fitted the data. To achieve this, we systematically restricted the full model, comparing the goodness of fit using likelihood ratio tests. Finally, in the third step, we inspected the ANOVA table of the final model and tested specific hypotheses about possible main effects or interactions. Kenward-Roger approximations to the degrees of freedom were used to adjust for small sample sizes (Kenward & Roger, 1997). When an interaction effect was significant, it was further investigated with follow-up contrast analyses.

## 3. Results

### 3.1. Intensity of the tactile stimulation

The mean intensities were not significantly different between the left and the right hand nor for the control participants (left hand: 0.53 ± 0.07 Watt, right hand: 0.50 ± 0.07 Watt; t(36) = 1.80, p = 0.08), nor for the FM patients (left hand: 0.51 ± 0.09 Watt, right hand: 0.48 ± 0.08 Watt; t(37) = 1.94, p = 0.06). There was no significant difference in intensities between control participants and FM patients (left hand: t(67.70) = 1.10, p = 0.27; right hand: t(72.40) = 1.44, p = 0.15).

The average self-reported intensities did not differ significantly for the left and the right hand, nor for the control participants (left hand: 3.08 ±1.71; right hand: 3.28 ± 1.74; t(36) = −1.97, p = 0.06), nor for the FM patients (left hand: 3.45 ±1.38; right hand: 3.62 ± 1.48; t(37) = −1.31, p = 0.20). There was no significant difference in self-reported intensities between controls and FM patients (left hand: t(69.07) = −1.05, p = 0.30; right hand: t(70.63) = −0.91, p = 0.36).^1^

### 3.2. Perceived threat value visual stimuli

Mean perceived threat scores were overall low, and were not significantly different when the lights were approaching (control participants: M = 0.51, SD = 1.33; FM patients: M = 1.05, SD = 2.20), compared to when they were receding (control participants: M = 0.41, SD = 1.09; FM patients: M = 1.02, SD = 2.22) from the participants (control participants: Z = 3, p = 0.37; FM patients: Z = 8.5, p = 0.89). Moreover, there was no significant difference in perceived threat scores between healthy controls and FM patients (Approaching: Z = 639, p = 0.36; Receding: Z = 635.5, p = 0.33).

### 3.3. Accuracy

All control participants had more than 80% correct task performance. Mean accuracy for this group was 97% (± 3%). Two FM patients had less than 80% correct (79.91% and 50.22%) and were excluded from further analyses. Mean accuracy for the remaining patients was 97% (± 2%). Accuracies were not further analyzed.

### 3.3. Reaction times

#### 3.3.1. Effect of visual stimulus direction

##### 3.3.1.1. Control participants

For control participants, the relationship between the RTs and the tactile targets, the different temporal delays at which the tactile stimuli were administered (from T1 to T7), the direction of the visual stimulus (approaching vs. receding) and the congruency of the tactile stimulation (congruent vs. incongruent to the visual stimuli) are represented in Figure 2.

**Figure 2.**
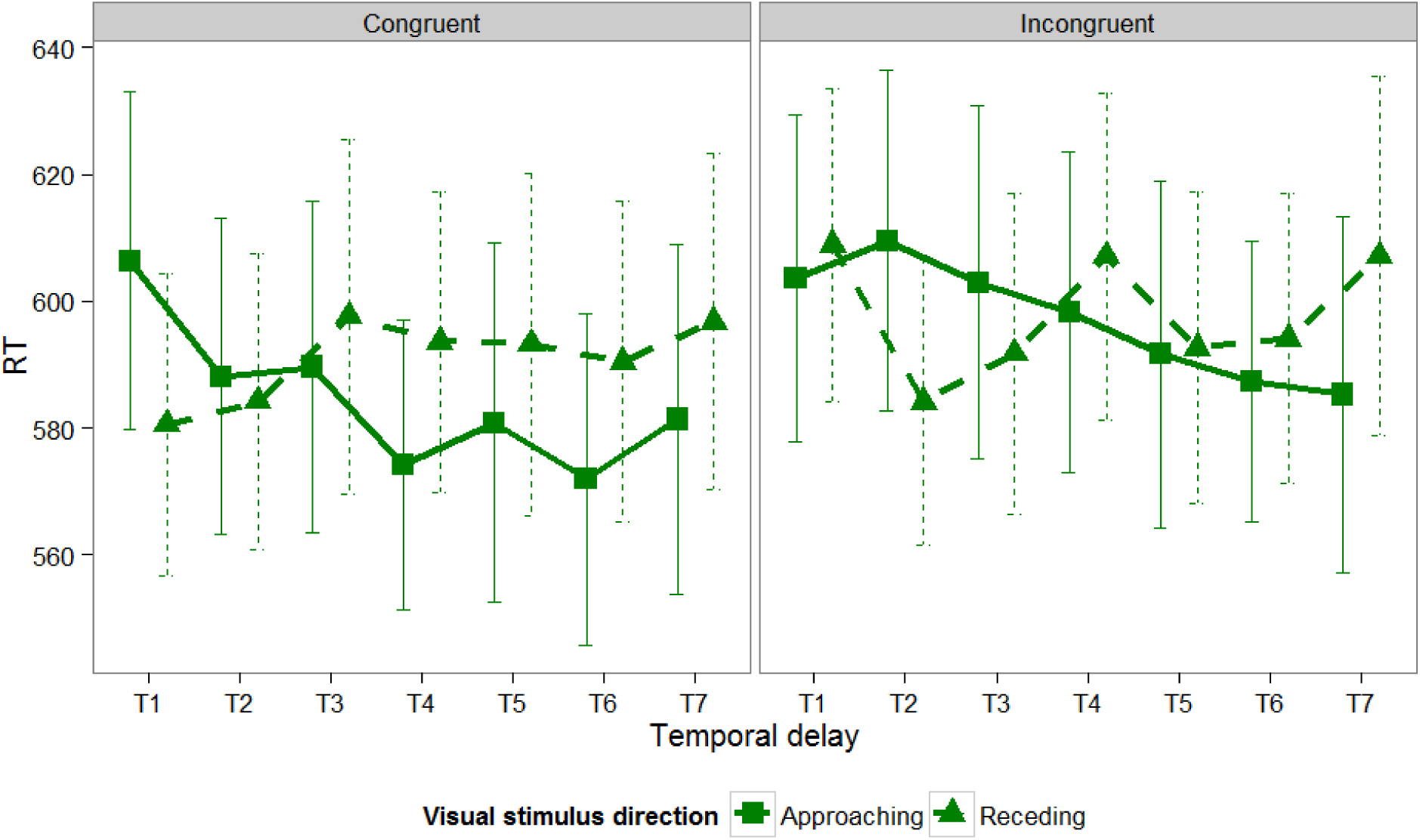
Mean RTs to the nociceptive targets and their associated standard errors in function of the different temporal delays at which the tactile stimuli were administered (from T1 to T7), the direction of the visual stimulus (approaching vs. receding) and the congruency of the tactile stimulation (congruent vs. incongruent to the visual stimulus) for control participants. The left side of the figure represents the congruent trials, while the right side represents the incongruent trials. The solid and dashed green lines depict the RTs in function of the temporal delays for the approaching and receding lights respectively.

The linear mixed effects model that demonstrated the best fit with the data, included all fixed factors together with their two-and three-way interactions, and a random subject-based intercept. The parameter estimates of the fixed effects together with their 95% confidence interval (CI) are shown in the Appendix (Table 1). In this final model, there was a significant main effect of visual stimulus direction (F(1,972) = 5.67; p = 0.02) and temporal delay (F(6,972) = 3.89; p < 0.001). Moreover, there was a significant interaction effect between visual stimulus direction and congruency (F(1,972) = 5.12; p = 0.02) and between visual stimulus direction and temporal delay (F(6,972) = 3.41; p = 0.002). Finally, the three-way interaction between visual stimulus direction, congruency and temporal delay was marginally significant (F(6,972) = 1.88; p = 0.08). The main effect of congruency (F(1,972) = 0.02; p = 0.90) and the interaction effect between congruency and temporal delay (F(6,972) = 1.25; p = 0.28) were not significant. To further investigate the three-way interaction and the two-way interactions, two separate linear mixed effect models were fitted for congruent and incongruent trials with visual stimulus direction and temporal delay as independent variables and RT as dependent variable.

For congruent trials, the model that demonstrated the best fit with the data included all fixed effect factors and their interaction, and a random subject-based intercept. In this model there was a significant main effect of visual stimulus direction (F(1,468) = 5.19; p = 0.02), a significant main effect of temporal delay (F(6,468) = 3.56; p = 0.002) and a significant interaction effect between visual stimulus direction and temporal delay (F(6,468) = 3.12; p = 0.005). Follow-up tests indicated that RTs at T1 were significantly faster for trials with receding visual stimuli than for those with approaching visual stimuli (χ2(1) = 5.19; p = 0.02). At T2 and T3 there was no significant difference in RTs between trials with approaching and receding visual stimuli (all χ2 < 0.75; all p > 0.35). Finally, from T4 to T7 the effect was reversed and RTs were now significantly faster for trials with approaching stimuli than for receding stimuli (T4: χ2(1) = 4.43; p = 0.04; T5: χ2(1) = 4.13; p = 0.04; T6: χ2(1) = 5.78; p = 0.02; T7: χ2(1) = 4.11; p = 0.04). Pairwise comparisons between the different temporal delays only revealed a marginally significant difference between T1 and T6 (t(36) = −1.54; p = 0.07) for approaching visual stimuli, indicating that, when lights were approaching, RTs were significantly slower at T1 than at T6. No other comparisons were significant nor for approaching, nor for receding visual stimuli (all |t| < 1.35; all p > 0.10).

For incongruent trials, a similar model was fitted to the data, that included all fixed effect factors and their interaction, and a random subject-based intercept. There was a significant main effect of temporal delay (F(6,468) = 3.59; p = 0.002), and a significant interaction effect between visual stimulus direction and temporal delay (F(6,468) = 2.66; p = 0.01). The main effect of visual stimulus direction did not reach significance (F(1,468) = 0.75; p = 0.39). Follow-up tests indicated that at T2, RTs for trials with receding visual stimuli were significantly faster than for those with approaching visual stimuli (χ2(1) = 5.45; p = 0.02). Conversely, at T7, RTs were significantly faster for trials with approaching visual stimuli than for trials with receding visual stimuli (χ2(1) = 9.70; p = 0.002). At the remaining temporal delays there was no significant difference between RTs for approaching versus receding stimuli trials (all χ2 < 0.75; all p > 0.35). Pairwise comparisons between the different temporal delays revealed a marginally significant difference between T2 and T7 (t(36) = −1.42; p = 0.08) for approaching visual stimuli. No other comparisons were significant, nor for approaching, nor for receding visual stimuli (all |t| < 1.02; p > 0.16).

##### 3.3.1.1. FM patients

For FM patients, the relationship between the RTs and the tactile targets, the different temporal delays at which the tactile stimuli were administered (from T1 to T7), the direction of the visual stimulus (approaching vs. receding) and the congruency of the tactile stimulation (congruent vs. incongruent to the visual stimulus) are represented in Figure 3.

**Figure 3.**
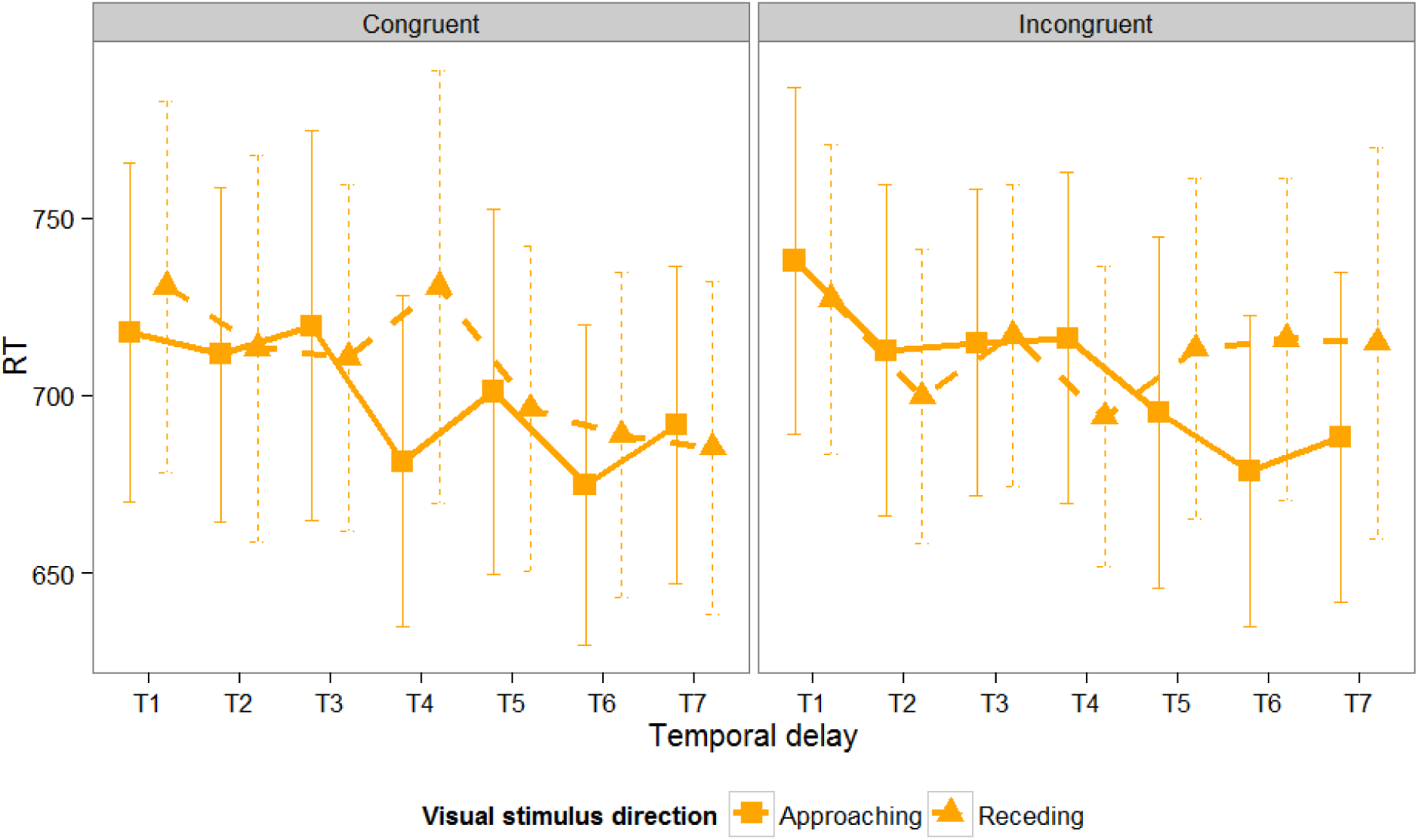
Mean RTs to the nociceptive targets and their associated standard errors in function of the different temporal delays at which the tactile stimuli were administered (from T1 to T7), the direction of the visual stimulus (approaching vs. receding) and the congruency of the tactile stimulation (congruent vs. incongruent to the visual stimulus) for FM patients. The left side of the figure represents the congruent trials, while the right side represents the incongruent trials. The solid and dashed orange lines depict the RTs in function of the temporal delays for the approaching and receding lights respectively.

The linear mixed effects model that demonstrated the best fit with the data, included all fixed factors together with their two-and three-way interactions, a random subject-based intercept, and a random effect for congruency and temporal delay. The parameter estimates of the fixed effects together with their 95% confidence interval (CI) are shown in the Appendix (Table 2). In this final model, there was a significant main effect of temporal delay (F(6,255.28) = 4.50; p < 0.001), a significant interaction between visual stimulus direction and temporal delay (F(6,740) = 2.70; p = 0.01), and a significant three-way interaction between visual stimulus direction, congruency and temporal delay (F(6,740) = 2.33; p = 0.03). The main effects of visual stimulus direction (F(1,740) = 0.26; p = 0.61) and congruency (F(1,568.41) = 1.45; p = 0.23) as well as the interaction effects of visual stimulus direction with congruency (F(1,740) = 0.62; p = 0.43) and of congruency with temporal delay (F(6,740) = 1.25; p = 0.28) did not reach significance. To further investigate the three-way interaction, two separate linear mixed effects models were fitted for congruent and incongruent trials with visual stimulus direction and temporal delay as independent variables and RT as dependent variable.

For congruent trials, the model that demonstrated the best fit with the data included the fixed factors and their interaction, and a random subject-based intercept. In this model there was a significant main effect of temporal delay (F(6,481) = 5.79; p < 0.001) and a significant interaction effect between visual stimulus direction and temporal delay (F(6,481) = 2.26; p = 0.04). The main effect of visual stimulus direction was not significant (F(1,481) = 0.22; p = 0.64). Follow-up tests indicated that RTs did not differ significantly between trials with approaching and receding visual stimuli from T1 to T3 (all χ2 < 1.20; all p > 0.25). At T4 and T6 RTs were (marginally) significantly faster for trials with approaching than for those with receding visual stimuli (T4: χ2(1) = 9.75; p = 0.002; T6: χ2(1) = 3.34, p = 0.07). At T5 and T7 RTs were not significantly different for trials with approaching and receding visual stimuli (all χ2 < 0.30; all p > 0.50). Pairwise comparisons between the different temporal delays showed that there were no significant differences in RTs between the temporal delays nor for approaching, nor for receding visual stimuli (all |t |< 0.93; all p > 0.14).

For incongruent trials, a similar model was fitted with all fixed factors, and their interaction, and a random subject-based intercept. In this model there was only a main effect of temporal delay (F(6,481) = 5.64; p < 0.001). The main effect of visual stimulus direction (F(1,481) = 0.31; p = 0.58) and the interaction effect between temporal delay and visual stimulus direction (F(6,481) = 1.57; p = 0.15) were not significant, indicating that RTs in this condition were less affected by the position of the visual stimuli in space.

#### 3.3.2. Effect of Group

The present study, as well as previous studies (Canzoneri et al., 2012; De Paepe et al., under review; Teneggi et al., 2013) suggest that stimuli approaching the stimulated hand have a larger spatially dependent effect on somatosensory processing compared to receding stimuli, or stimuli approaching the opposite side. Therefore, to assess potential differences in the spatially dependent effect of visual stimuli on somatosensory processing between FM patients and control participants we only used the congruent trials with approaching visual stimuli.

The model that demonstrated the best fit with the data included a main effect of temporal delay, a main effect of group, and a random subject-based intercept. Adding the interaction effect between temporal delay and group did not significantly increase the fit of the model. Therefore, the interaction effect was not included in the model. In the final model, there was a significant main effect of temporal delay (F(6,444) = 8.91; p < 0.001), and a significant main effect of group (F(1,73) = 8.53; p = 0.005), indicating that FM patients responded significantly slower than the control participants.

Finally, to assess whether the curves describing the relationship between the RTs and the temporal delay were different between FM patients and healthy controls, we simplified the model for both groups by considering temporal delay as a continuous variable instead of a factor, so that T1 corresponds to 170 ms, T2 to 450 ms, T3 to 730 ms, T4 to 1010 ms, T5 to 1290 ms, T6 to 1570 ms and T7 to 1850 ms. The nature of the relationship between the independent variable temporal delay and the dependent variable RT was investigated by fitting models with RT (on the original scale) as dependent variable and temporal delay as independent variable separately for FM patients and control participants^2^. At each time the restricted models (with temporal delay as continuous variable) were compared with the full model (with temporal delay as categorical variable). For the control participants a linear relationship was considered assuming a constant increase/decrease of RTs as a function of temporal delay. This model did not fit the data significantly worse than the model with temporal delay as a categorical variable (χ2(5) = 5.51, p = 0.36). For control participants the relationship between the RTs and temporal delay could thus be adequately described by assuming a linear model. For FM patients the same strategy was applied. First, a linear relationship was considered, assuming a constant increase/decrease of RTs as a function of the temporal delay. This model fitted the data significantly worse than the model with temporal delay as a categorical predictor (χ2(5) = 12.05; p = 0.03). Next, a quadratic relationship was considered by adding the square of the independent variable temporal delay to the model. This model did not fit the data significantly worse than the full model (χ2(4) = 8.41, p = 0.08). For FM patients the relationship between the RTs and temporal delay could be adequately described by assuming a quadratic model. The predicted values of these models, centered around the group means, together with their fitted curves are shown in Figure 4.

**Figure 4.**
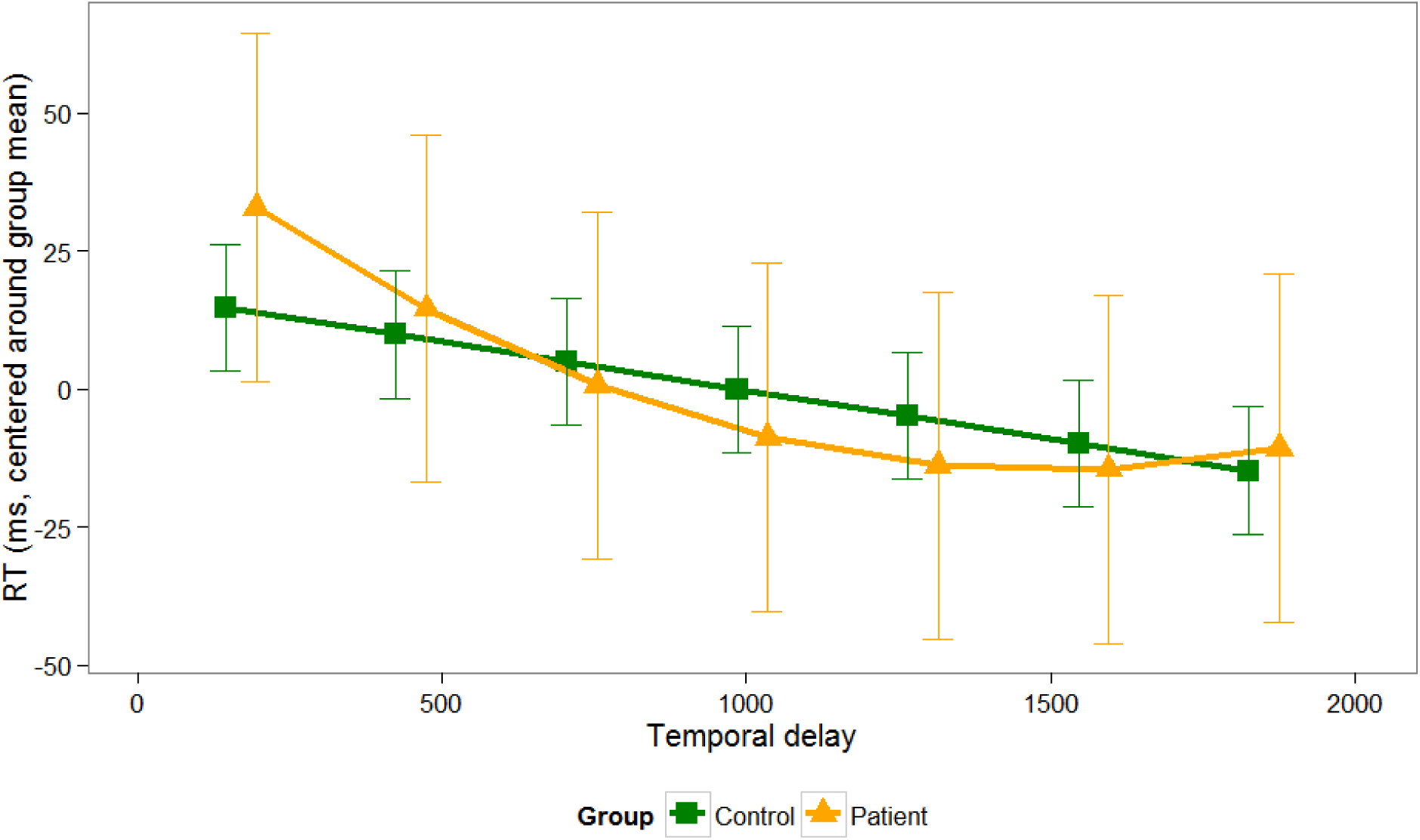
Predicted values and standard errors for the relationship between RTs to the tactile targets in function of the different temporal delays at which the tactile stimuli were administered (from T1 to T7), and the group to which participants belong (FM patient vs. control). To easily compare the curves for both groups, the predicted values are centered around the group mean. For FM patients a quadratic model fitted the data best (orange curve), whereas for control participants, a linear model adequately fitted the data (green curve).

## 4. Discussion

In the present study, we investigated whether attention towards stimuli approaching the participant’s body differed for FM patients compared to healthy control participants. To this end, the differential influence of dynamical visual stimuli on tactile processing was studied for control participants and FM patients. For control participants, we found that visual stimuli presented near the stimulated hand influenced tactile processing more than visual stimuli presented far from the hand. For FM patients this difference was less clear. The influence of the approaching visual stimuli on tactile processing along a continuous spatial range (from near to far) was compared for controls and FM patients. Although there were no significant differences in RTs between both groups, the curves describing the RTs along the spatial range were different for FM patients compared to healthy control participants. However, as this difference was only found when curve-fitting the data, our results should be interpreted with caution and need further corroboration and replication.

The ability to interact with the world closely surrounding us depends on the integration of somatosensory stimuli on the body space, external stimuli presented near the body, and information about body posture (Cardinali, Brozzoli, & Farnè, 2009; Rizzolatti, Scandolara, Matelli, & Gentilucci, 1981; Spence & Driver, 2004). For visuo-tactile integration it has been demonstrated in monkeys that this ability relies on neurons with multimodal receptive fields (RFs), found mainly in the premotor and intraparietal areas (Graziano, Hu, & Gross, 1997; Graziano, Yap, & Gross, 1994). These neurons are activated in response to both tactile stimuli and to visual stimuli occurring close to the stimulated body parts. Graziano & Cooke (2006) argued that one of the main functions of these cortical areas is to maintain a margin of safety around the body and to coordinate defensive actions. In humans, the use of a peripersonal frame of reference for the localization of somatosensory stimuli has been demonstrated both in neuropsychological studies with patients suffering from crossmodal extinction consecutive to right hemisphere damages (di Pellegrino, Làdavas, & Farnè, 1997; Làdavas, di Pellegrino, Farnè, & Zeloni, 1998) and in healthy participants using crossmodal congruency tasks (Holmes, Sanabria, Calvert, & Spence, 2006; Sambo & Forster, 2009; Spence, Pavani, & Driver, 2000, 2004). Neuroimaging studies provide support for the role of the frontal cortex (Lloyd, Shore, Spence, & Calvert, 2003) and the parietal cortex (Makin, Holmes, & Zohary, 2007) in the multisensory representation of the body. This fronto-parietal network might in turn boost the activity of unisensory areas, facilitating the processing of sensory inputs from each modality (Kennett, Eimer, Spence, & Driver, 2001; Macaluso, Frith, & Driver, 2000; Taylor-Clarke, Kennett, & Haggard, 2002). Overall, these studies show that the crossmodal influence of visual stimuli on tactile processing is more pronounced when visual stimuli are presented near the stimulated body limb as opposed to far from that limb. Recently, these results were extended to the crossmodal interaction between visual stimuli and stimuli specifically activating nociceptive skin receptors (De Paepe et al., 2015, 2014, in preparation).

The studies mentioned so far have investigated multimodal integration in the peripersonal space using static stimuli at two fixed positions (near vs. far). Some studies have also demonstrated this phenomenon with moving stimuli (e.g., Canzoneri et al., 2012; Van der Biest, Legrain, De Paepe, & Crombez, 2015; De Paepe et al., under review). The use of moving stimuli is more ecologically valid and more comparable to animal studies investigating multimodal integration in the peripersonal space (Dong, Chudler, Sugiyama, Roberts, & Hayashi, 1994; Duhamel, Colby, & Goldberg, 1998; Graziano et al., 1997). Furthermore, studies in both humans and monkeys (Graziano et al., 1997; Makin et al., 2007) have shown that the neural systems representing the peripersonal space show a preference for moving stimuli. The use of moving stimuli also enables the investigation of the influence of external stimuli along a continuous spatial range (from near to far). For example, Canzoneri et al., (2012) measured reaction times (RTs) to a tactile stimulus applied to the right index finger while dynamical sounds, which gave the impression of either approaching or receding from the subject’s hand, were presented. Tactile stimuli were delivered at different temporal delays from the onset of the sound, such that it occurred when the sound source was perceived at varying distances from the body. Participants were asked to respond as fast as possible, trying to ignore the sound. They found that an auditory stimulus speeded up the processing of a tactile stimulus applied to the hand when the sound was administered within a limited distance from the hand. Moreover, results suggested that approaching sounds had a stronger spatially dependent effect on tactile processing compared to receding sounds. Recently, we adapted this paradigm to investigate the influence of dynamical visual stimuli on nociceptive processing in healthy volunteers (De Paepe et al., under review). In this study, a visual stimulus was either approaching or receding the participant’s left or right hand on each trial. At different temporal delays from the onset of the visual stimulus, a nociceptive stimulus was applied either at the same or the opposite hand, so that it was presented when the visual stimulus was perceived at varying distances from the hand. Participants were asked to respond as fast as possible at which side they perceived a nociceptive stimulus. We found similar results as Canzoneri et al. (2012): RTs were fastest when the visual stimulus appeared near the stimulated hand and approaching lights had a stronger spatially dependent effect on nociceptive processing, compared to receding lights.

In the present study we were able to replicate these results in healthy, community dwelling participants using the same paradigm with tactile stimuli instead of nociceptive stimuli. Results showed that at low temporal delays, RTs were faster for receding visual stimuli, whereas at high temporal delays, RTs were faster for approaching visual stimuli. This indicates that the visual stimuli had the largest impact on tactile processing when they were presented near the participants’ hand. In contrast with the previous study (De Paepe et al., under review) we found that the effect of the temporal delay on RTs was dependent on the visual stimulus direction both for congruent and incongruent visual stimuli. However, further investigation of these interactions showed that for congruent visual stimuli there was a clear pattern going from faster RTs for receding visual stimuli trials at low temporal delays, over no difference at intermediate delays, to faster RTs for approaching visual stimuli at high temporal delays. Conversely, for incongruent trials there was only a significant difference between approaching and receding trials at T2 and T7, probably indicating that the effect of the visual stimuli was less robust when visual stimuli were approaching or receding the opposite (non-stimulated) hand. Our results are in line with studies in monkeys and humans, suggesting that the representation of peripersonal space is body-part centered (De Paepe et al., under review; di Pellegrino et al., 1997; Graziano et al., 1997).

Next, we were interested whether a similar response pattern could be found in FM patients. In accordance with the previous study (De Paepe et al., under review), we found that the direction of the visual stimuli only influenced the relationship between the temporal delay and the RTs for congruent trials and not for incongruent trials. Further investigation of the congruent trials showed that RTs were faster for approaching versus receding visual stimulus trials at some of the high temporal delays (T4 and T6). At low temporal delays there was no significant difference. This may indicate that FM patients already had faster RTs for approaching stimuli at low temporal delays, causing the difference between receding and approaching trials to disappear. This would be in agreement with the idea that FM patients have altered reactions to stimuli at further distance from the body compared to healthy control participants. However, it should be noted that there was a large variability in RTs between patients: The variance of the random intercept explained 32% of the total variance for the FM data, compared to only 5% for the data of the control participants. This huge amount of variance between patients may have created a lot of noise, masking significant differences within the data. Therefore, caution is warranted in interpreting our results, and we feel yet unable to draw strong inferences. Further research may try to reduce the variability amongst patients, by categorizing patients into more similar groups based on age, onset of FM, amount of pain, or individual difference variables such as anxiety and catastrophizing that may be considered to play an important role.

Paradigms similar to the one of Canzoneri et al. (2012) have also been used to demonstrate modulations of the peripersonal space (Taffou & Viaud-Delmon, 2014; Teneggi et al., 2013), comparing the influence of approaching stimuli on tactile processing across different experimental conditions. In the present study, we compared the influence of the visual stimuli, approaching the stimulated hand of the participants, on tactile processing between control participants and FM patients. The comparison between both groups was only made for trials in which the visual stimulus was approaching the hand that received the tactile stimulus, because results of the present and previous studies indicated that stimuli approaching the stimulated hand have a larger spatially dependent effect on somatosensory processing than receding stimuli or stimuli approaching the opposite hand (Canzoneri et al., 2012; De Paepe et al., under review; Teneggi et al., 2013). Moreover, previous research investigating modulations of the peripersonal space across different conditions or groups also focused exclusively on approaching stimuli. Therefore, to increase the power of the model and for the sake of parsimony, we only considered these trials. Results showed that RTs decreased with increasing temporal delays, but this decrease was not significantly different for FM patients compared to control participants. Additionally, we found that FM patients responded slower than control participants. This is in accordance with previous studies (Correa, Miró, Martínez, Sánchez, & Lupiáñez, 2011; del Paso, Montoro, & Duschek, 2015) and was suggested to be associated with a reduced capacity to maintain the level of activation necessary to perform the task (Correa et al., 2011; Miró et al., 2011), as shown by studies demonstrating impairments in the speed of processing (C-oté & Moldofsky, 1997), in sustained attention (Dick, Eccleston, & Crombez, 2002; Dick, Verrier, Harker, & Rashiq, 2008) and in the alerting system (Miró et al., 2011) in FM patients. The fact that we did not find any differences between FM patients and healthy controls can be attributed to a number of factors. First, the cognitive deficits identified in FM patients, mentioned above, may make it difficult to find significant results in a cognitive demanding task such as the present study. Moreover, this may also explain the large inter-individual variability in RTs found in this study. Second, the large inter-individual variability in the FM group may have masked an effect in this group, making it difficult to find any differences between groups. Third, it should be noted that not all studies found evidence for perceptual amplification in modalities other than pain in FM patients (Carrillo-de-la-Peña, Triñanes, González-Villar, Gómez-Perretta, & García-Larrea, 2014; Lorenz, 1998; Van Damme et al., 2015) and the studies that did find differences used auditory instead of visual stimuli (Dohrenbusch, Sodhi, Lamprecht, & Genth, 1997; Hollins et al., 2009; McDermid et al., 1996).

Finally, we assessed differences in the spatially-dependent effect of the visual stimuli on tactile processing by fitting curves to the RTs at the different temporal delays for both groups. For control participants a linear function adequately described the data, indicating that RTs showed a constant decrease with increasing temporal delays. This is in contrast with the previous study in which a quadratic function (De Paepe et al., under review) described the data best. However, in the previous study nociceptive targets were used instead of tactile targets. A possible explanation could be that in the context of nociceptive stimuli the visual stimuli approaching the participants are experienced as more threatening than in the context of tactile stimuli. Indeed, although the perceived threat value of the approaching lights was low in both studies, it was significantly higher in the previous study (M = 1.78, SD = 2.47) (De Paepe et al., under review), than in the present study (M = 0.51, SD = 1.33), t(36.90) = 2.42; p = 0.02. Previous studies have shown that the spatially-dependent effect of external stimuli on nociceptive and tactile processing is stronger for approaching than for receding stimuli (Ball & Tronick, 1971; Bremmer, Duhamel, Ben Hamed, & Graf, 2002; Canzoneri et al., 2012; Colby, Duhamel, & Goldberg, 1993; De Paepe et al., under review; Duhamel, Bremmer, Benhamed, & Graf, 1997; Kandula, Hofman, & Dijkerman, 2014; Schiff, Caviness, & Gibson, 1962; Schiff, 1965; Tinbergen, 1951). It could be that this is especially true in a threatening context, in which it is crucial to quickly prepare an appropriate defensive response. This is consistent with research showing that individuals underestimate the time a visual stimulus approaching them will collide with them when the stimulus is threatening (snakes, spiders, angry faces) compared to when it is non-threatening (butterflies, rabbits, neutral faces) (Brendel, DeLucia, Hecht, Stacy, & Larsen, 2012; Vagnoni, Lourenco, & Longo, 2012). Although an intriguing hypothesis, future studies need to address this issue directly, by comparing the spatially-dependent effect of visual stimuli on nociceptive and tactile processing, preferably within the same participants. Conversely, for FM patients we found that a quadratic function best described the data. RTs showed a steep decrease at low temporal delays, to stabilize at higher temporal delays. The perceived threat value of the approaching stimuli for the patients was comparable to that of the previous study (De Paepe et al., under review) (M = 1.05, SD =2.20; t(52.05) = 1.22, p = 0.23) and was somewhat higher (although not significantly) than the perceived threat value for the control group. This could explain why in agreement with the previous study, a quadratic function fitted the data best for FM patients, while a linear function fitted the data best for the healthy control participants. The fact that FM patients showed a quicker decline in RTs at high temporal delays seems compatible with the hypothesis that FM patients would have a heightened attention for stimuli entering the peripersonal space. The difference between FM patients and healthy control participants, could result from the fact that FM patients are ‘hypervigilant’ for sensory information. The mechanisms of hypervigilance or still not completely understood. Although different theories exist (e.g., Chapman, 1978; Hollins et al., 2009; McDermid et al., 1996; Rollman & Lautenbacher, 1993), the ‘attentional gain control theory’ of Hollins et al. (2009) seems most compatible with the present results. Hollins et al. (2009) proposed that “hypervigilance begins as a cognitive process, in which an individual is concerned about, and therefore closely monitors, particular types of sensations – especially those that, while not necessarily unpleasant in themselves, accompany or warn impending pain. (…) Sustained direction of this affect-charged attention to a particular kind of stimulation, produces, over time, an increase in the perceptual gain for all stimuli of that type” (Hollins et al., 2009, p 221). Because of this heightened attention for stimuli signaling potential threat to the body, FM patients may show a stronger spatially dependent effect of the approaching lights on tactile reaction times than control participants.

It should be noted that based on the present study we cannot be sure whether any differences observed between controls and FM patients are due to a modulation of the peripersonal space (i.e. whether the same system is now used to perceive stimuli both at near and far positions) or rather to a more extreme reaction to stimuli in the extrapersonal space. In the latter view, two distinct systems would still be distinguishable to perceive stimuli at a proximal versus farther positions, but stimuli at farther positions are now perceived equally threatening or relevant as proximal stimuli. This contrast is difficult to make and depends on the conceptualization of the peripersonal space. Here we defined peripersonal space as ‘the space in which stimuli on the body space are integrated with stimuli occurring in the external world’. In some of the previous studies this integration seems to be best described by a sigmoid function (Canzoneri et al., 2012; Taffou & Viaud-Delmon, 2014; Teneggi et al., 2013), and the abscissa at the inflection point of the sigmoidal curve is taken as the boundary of the peripersonal space. This boundary between peripersonal and extrapersonal space seems rather arbitrary, and in the present study sigmoid functions did not fit our data adequately. We argue that a strict boundary between the peripersonal and extrapersonal space is unlikely to exist, and that some multimodal integration will probably also occur in what we call the ‘extrapersonal’ space (see e.g., De Paepe et al., 2015, 2014; Rizzolatti et al., 1981). Therefore, ‘a modulation of the peripersonal space’ should be seen as a difference in attention towards stimuli approaching the body, rather than the extension or reduction of a strict boundary between peripersonal and extrapersonal space.

This study has some limitations. First of all, the high inter-individual variability in the patient data posed a great challenge for analyzing and interpreting the data. As mentioned above, this could be due to attentional and information processing problems in FM patients (Correa et al., 2011; Miró et al., 2011) and may always be a problem when using RT as dependent variable. Future studies should focus on paradigms using different measures (such as accuracy) (e.g., temporal order judgment tasks). Second, the present study was not designed to explain the underlying mechanisms of possible modulations of the peripersonal space due to FM. One possibility is that FM patients appraise bodily sensations as dangerous and are therefore more likely to continuously scan the body and the space closely surrounding it for threatening sensations (Chapman, 1978; Crombez, Eccleston, Van den Broeck, Goubert, & Van Houdenhove, 2004). This could lead to the perceptual amplification and possibly also to a modulation of the peripersonal space compared to healthy control participants. In this respect it could be interesting to take psychological variables such as catastrophizing about pain or body vigilance into account. Apart from adding these variables to the model as covariates, another possibility would be to categorize the participants into several subgroups based on these variables. As mentioned above, this could have the additional advantage of reducing inter-individual variability. It is possible that only a particular subgroup of patients showed the effect. Unfortunately, our study lacked statistical power for such an approach. Third, in the present study we investigated the influence of visual stimuli on tactile processing. Previous studies showing evidence for perceptual amplification have largely used auditory stimuli (Dohrenbusch et al., 1997; Hollins et al., 2009; McDermid et al., 1996). It could be that FM patients do not show perceptual amplification for visual stimuli, or that perceptual amplification is much weaker for visual stimuli. Moreover, studies showing modulations of the peripersonal space have used auditory stimuli in a detection task, in which participants have to respond as quickly as possible when they felt something (Taffou & Viaud-Delmon, 2014; Teneggi et al., 2013). In the present study we used a localization task, in which participants have to respond as quickly as possible which hand was stimulated. To be able to compare our results with those of previous studies, it could be interesting to adapt the paradigm, using auditory stimuli in a detection task instead of visual stimuli in a localization task.

## Data Availability

All data produced in the present study are available upon reasonable request to the authors

## 5. Acknowledgments

A.L. De Paepe is supported by the Research Foundation Flanders (FWO, Belgium). V. Legrain is supported by the Fund for Scientific Research of the French speaking Community of Belgium (F.R.S.-FNRS).

## Appendix

**Table 1.**
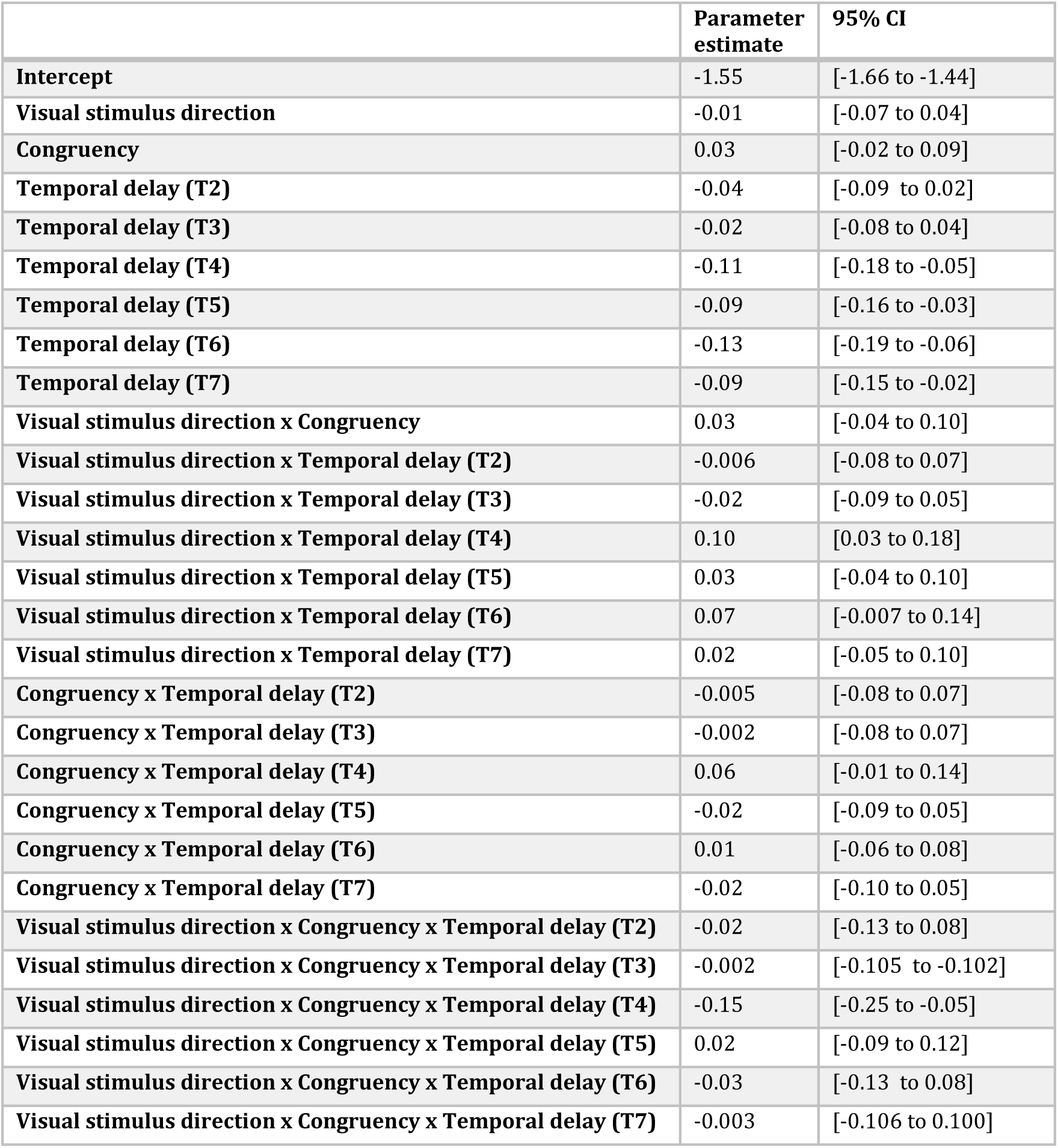
Parameter estimates and associated 95% confidence intervals (CI) for the fixed effects of the fitted model predicting the inverse transformed RTs in function of visual stimulus direction, congruency and temporal delay for FM patients.

|  | **Parameter estimate** | **95% CI** |
|---|---|---|
| **Intercept** | -1.55 | [-1.66 to -1.44] |
| **Visual stimulus direction** | -0.01 | [-0.07 to 0.04] |
| **Congruency** | 0.03 | [-0.02 to 0.09] |
| **Temporal delay (T2)** | -0.04 | [-0.09 to 0.02] |
| **Temporal delay (T3)** | -0.02 | [-0.08 to 0.04] |
| **Temporal delay (T4)** | -0.11 | [-0.18 to -0.05] |
| **Temporal delay (T5)** | -0.09 | [-0.16 to -0.03] |
| **Temporal delay (T6)** | -0.13 | [-0.19 to -0.06] |
| **Temporal delay (T7)** | -0.09 | [-0.15 to -0.02] |
| **Visual stimulus direction x Congruency** | 0.03 | [-0.04 to 0.10] |
| **Visual stimulus direction x Temporal delay (T2)** | -0.006 | [-0.08 to 0.07] |
| **Visual stimulus direction x Temporal delay (T3)** | -0.02 | [-0.09 to 0.05] |
| **Visual stimulus direction x Temporal delay (T4)** | 0.10 | [0.03 to 0.18] |
| **Visual stimulus direction x Temporal delay (T5)** | 0.03 | [-0.04 to 0.10] |
| **Visual stimulus direction x Temporal delay (T6)** | 0.07 | [-0.007 to 0.14] |
| **Visual stimulus direction x Temporal delay (T7)** | 0.02 | [-0.05 to 0.10] |
| **Congruency x Temporal delay (T2)** | -0.005 | [-0.08 to 0.07] |
| **Congruency x Temporal delay (T3)** | -0.002 | [-0.08 to 0.07] |
| **Congruency x Temporal delay (T4)** | 0.06 | [-0.01 to 0.14] |
| **Congruency x Temporal delay (T5)** | -0.02 | [-0.09 to 0.05] |
| **Congruency x Temporal delay (T6)** | 0.01 | [-0.06 to 0.08] |
| **Congruency x Temporal delay (T7)** | -0.02 | [-0.10 to 0.05] |
| **Visual stimulus direction x Congruency x Temporal delay (T2)** | -0.02 | [-0.13 to 0.08] |
| **Visual stimulus direction x Congruency x Temporal delay (T3)** | -0.002 | [-0.105 to -0.102] |
| **Visual stimulus direction x Congruency x Temporal delay (T4)** | -0.15 | [-0.25 to -0.05] |
| **Visual stimulus direction x Congruency x Temporal delay (T5)** | 0.02 | [-0.09 to 0.12] |
| **Visual stimulus direction x Congruency x Temporal delay (T6)** | -0.03 | [-0.13 to 0.08] |
| **Visual stimulus direction x Congruency x Temporal delay (T7)** | -0.003 | [-0.106 to 0.100] |

**Table 2.**
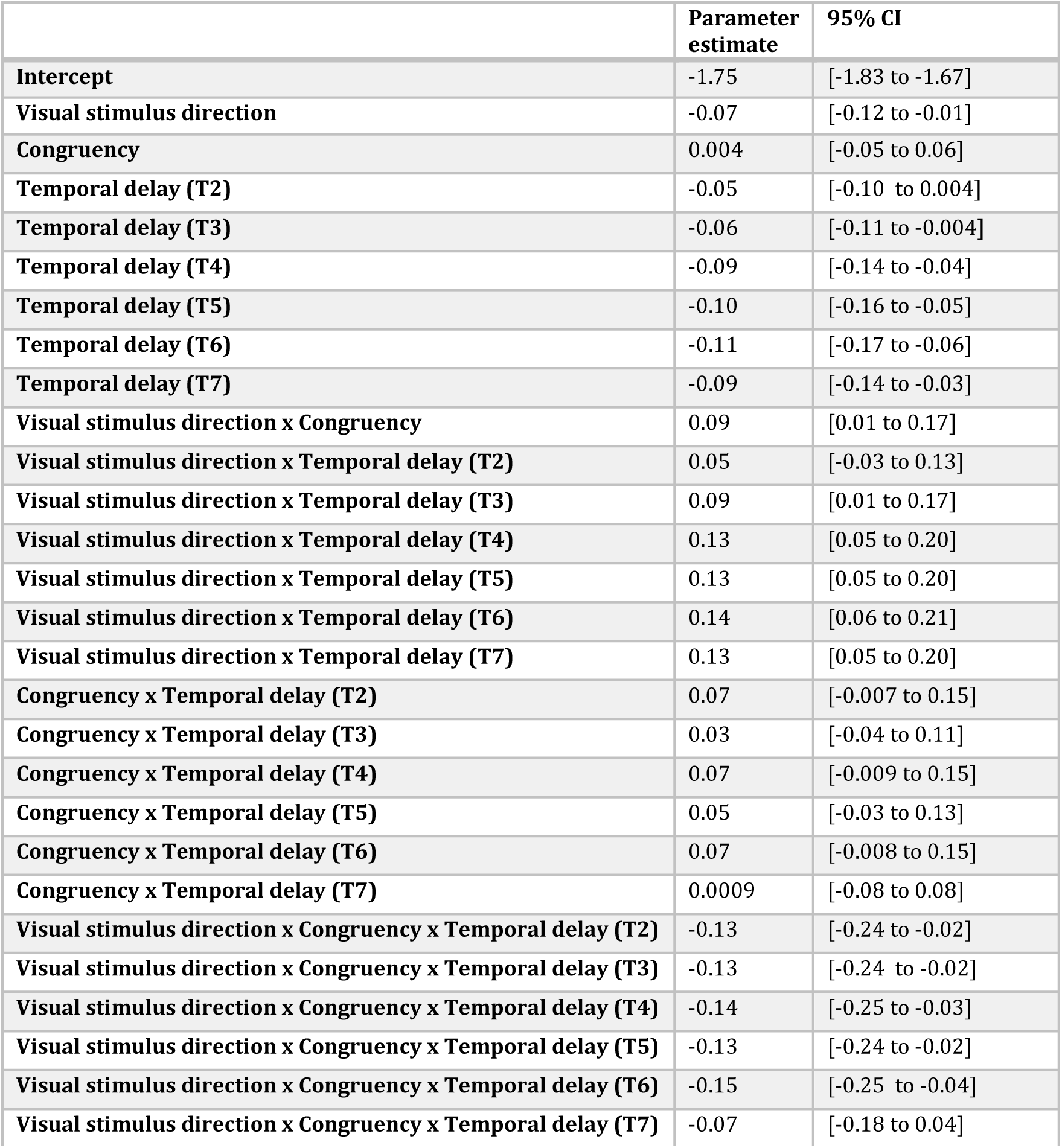
Parameter estimates and associated 95% confidence intervals (CI) for the fixed effects of the fitted model predicting the inverse transformed RTs in function of the visual stimulus direction, congruency and temporal delay for control participants.

## Footnotes

1 To control whether the side of the stimulation had an influence on the task performance, the *side of the tactile stimulus* was added to the model as additional variable. For control participants, there was a significant interaction effect between *congruency* and *side of stimulation* (F(1,2004) = 6.59; *p* = 0.01), showing that participants were significantly faster to respond on congruent as opposed to incongruent trials when their right hand was stimulated (χ^2^(1) = 23.39; *p* < 0.001), but not when their left hand was stimulated (χ^2^(1) = 1.46; *p* = 0.23). Nor the main effect of side of stimulation (F(1,2004) = 0.21; *p* = 0.65), nor any of its interactions with other variables were significant (all F < 1; *p* > 0.45). For FM patients, there was a marginally significant main effect of the *side of stimulation* (F(1,1998) = 2.82; *p* = 0.09), indicating that patients responded faster when the right hand was stimulated than when the left hand was stimulated. However, none of the interaction effects between side of stimulation and any of the other variables were significant, indicating that this did not influence the effect of the lights on somatosensory processing (all F < 0.6; *p* > 0.70). Therefore, the side of stimulation was left out of any further analyses.

2 It should be noted that previous studies investigating modulations of the peripersonal space fitted a sigmoid function of the following form to their data: 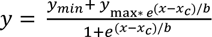, where *x* represents the independent variable (temporal delay), *y* the dependent variable (i.e., RT), *y_min_* and *y_max_* the lower and upper saturation levels of the sigmoid, *x_c_* the value of the abscissa at the central point of the sigmoid (i.e., the value of *x* at which *y = (y_min_+ y_max_)/2*) and *b* establishes the slope of the sigmoid at the central point (Teneggi et al., 2013; Taffou et al., 2014). For the sake of comparability, we tried to fit this sigmoid function to our data. Values of the parameters *y_min_* and *y_max_* were set a priori to the minimum and maximum values for each participant, and the estimated parameters were the central position of the sigmoid (*x_c_*) and the slope of the sigmoid at the central point. A sigmoid function could only be fitted for 21 out of 37 control participants (56%), and for 23 out of 38 FM patients (61%). For these participants the parameter *x_c_* was compared for control participants and FM patients as a measure of the distance, at which the visual stimuli started affecting tactile RTs. Results showed that this parameter was not significantly different for FM patients (*M* = 789.09, *SD* = 331.03) as compared to healthy control participants (*M* = 808.70; *SD* = 305.91) (t(41.99) = 0.20; *p* = 0.80). Since a sigmoid function could only be fitted to less than 60% of the data, this approach seems less appropriate for the present study.

## References

Arendt-Nielsen, L., & Henriksson, K. G. (2007). Pathophysiological mechanisms in chronic musculoskeletal pain (fibromyalgia): the role of central and peripheral sensitization and pain disinhibition. Best Practice & Research Clinical Rheumatology, 21(3), 465–480. 10.1016/j.berh.2007.03.007

Ball, W., & Tronick, E. (1971). Infant responses to impending collision - optical and real. Science, 171(3973), 818–&.

Bassolino, M., Serino, A., Ubaldi, S., & Làdavas, E. (2010). Everyday use of the computer mouse extends peripersonal space representation. Neuropsychologia, 48(3), 803–811. 10.1016/j.neuropsychologia.2009.11.009

Bremmer, F., Duhamel, J. R., Ben Hamed, S., & Graf, W. (2002). Heading encoding in the macaque ventral intraparietal area (VIP). European Journal of Neuroscience, 16, 1554–1568. 10.1046/j.1460-9568.2002.02207.x

Brendel, E., DeLucia, P. R., Hecht, H., Stacy, R. L., & Larsen, J. T. (2012). Threatening pictures induce shortened time-to-contact estimates. Attention, Perception, & Psychophysics, 74(5), 979–987. 10.3758/s13414-012-0285-0

Canzoneri, E., Magosso, E., & Serino, A. (2012). Dynamic Sounds Capture the Boundaries of Peripersonal Space Representation in Humans. PLoS ONE, 7(9), 3–10. 10.1371/journal.pone.0044306

Cardinali, L., Brozzoli, C., & Farnè, A. (2009). Peripersonal Space and Body Schema: Two Labels for the Same Concept? Brain Topography, 21(3-4), 252–260. 10.1007/s10548-009-0092-7

Carrillo-de-la-Peña, M. T., Triñanes, Y., González-Villar, a, Gómez-Perretta, C., & García-Larrea, L. (2014). Filtering out repetitive auditory stimuli in fibromyalgia: A study of P50 sensory gating. European Journal of Pain (London, England), 1–9. 10.1002/ejp.627

Chapman, C. R. (1978). The perception of noxious events. In R. A. Sternbach (Ed.), The Psychology of Pain (pp. 169–203). New York: Raven Press.

Colby, C. L., Duhamel, J. R., & Goldberg, M. E. (1993). Ventral intraparietal area of the macaque: Anatomic location and visual response properties. Journal of Neurophysiology, 69, 902–91vt4.

Correa, Á., Miró, E., Martínez, M. P., Sánchez, A. I., & Lupiáñez, J. (2011). Temporal preparation and inhibitory deficit in fibromyalgia syndrome. Brain and Cognition, 75(3), 211–216. 10.1016/j.bandc.2010.11.005

Coté, K. A., & Moldofsky, H. (1997). Sleep, daytime symptoms, and cognitive performance in patients with fibromyalgia. The Journal of Rheumatology, 24(10), 2014–2023.

Crombez, G., Eccleston, C., Van den Broeck, A., Goubert, L., & Van Houdenhove, B. (2004). Hypervigilance to Pain in Fibromyalgia. The Clinical Journal of Pain, 20(2), 98–102. 10.1097/00002508-200403000-00006

Crombez, G., Van Damme, S., & Eccleston, C. (2005). Hypervigilance to pain: An experimental and clinical analysis. Pain, 116, 4–7. 10.1016/j.pain.2005.03.035

Damme, S. Van, Crombez, G., Wiech, K., Legrain, V., Peters, M. L., & Eccleston, C. (2009). Why become more general when we can be more specific? Comment on Hollins et al. “Perceived intensity and unpleasantness of cutaneous and auditory stimuli: An evaluation of the generalized hypervigilance hypothesis” [Pain 2009;141:215-221], and on Rollman. Pain, 144(3), 342–343. 10.1016/j.pain.2009.04.035

De Paepe, A. L., Crombez, G., & Legrain, L. (n.d.). What’s coming near? The influence of dynamical visual stimuli on nociceptive processing. PLoS ONE.

De Paepe, A. L., Crombez, G., & Legrain, V. (2015). From a Somatotopic to a Spatiotopic Frame of Reference for the Localization of Nociceptive Stimuli. Plos One, 10(8), e0137120. 10.1371/journal.pone.0137120

De Paepe, A. L., Crombez, G., Spence, C., & Legrain, V. (2014). Mapping nociceptive stimuli in a peripersonal frame of reference: evidence from a temporal order judgment task. Neuropsychologia, 56, 219–228. 10.1016/j.neuropsychologia.2014.01.016

del Paso, G. A. R., Montoro, C. I., & Duschek, S. (2015). Reaction time, cerebral blood flow, and heart rate responses in fibromyalgia: Evidence of alterations in attentional control. Journal of Clinical and Experimental Neuropsychology, 37(4), 414–428. 10.1080/13803395.2015.1023265

di Pellegrino, G., Làdavas, E., & Farnè, A. (1997). Seeing where your hands are. Nature, 388(6644), 730. 10.1038/41921

Dick, B., Eccleston, C., & Crombez, G. (2002). Attentional functioning in fibromyalgia, rheumatoid arthritis, and musculoskeletal pain patients. Arthritis and Rheumatism, 47(6), 639–644. 10.1002/art.10800

Dick, B., Verrier, M. J., Harker, K. T., & Rashiq, S. (2008). Disruption of cognitive function in Fibromyalgia Syndrome. Pain, 139(3), 610–616. 10.1016/j.pain.2008.06.017

Dohrenbusch, R., Sodhi, H., Lamprecht, J., & Genth, E. (1997). Fibromyalgia as a disorder of perceptual organization? An analysis of acoustic stimulus processing in patients with widespread pain. Zeitschrift F{ü}r Rheumatologie, 56(6), 334–341. 10.1007/s003930050047

Dong, W. K., Chudler, E. H., Sugiyama, K., Roberts, V. J., & Hayashi, T. (1994). Somatosensory, multisensory, and task-related neurons in cortical area 7b (PF) of unanesthetized monkeys. Journal of Neurophysiology, 72(2), 542–564.

Duhamel, J. R., Bremmer, F., Benhamed, S., & Graf, W. (1997). Spatial invariance of visual receptive fields in parietal cortex neurons. Nature, 389(October), 845–848.

Duhamel, J. R., Colby, C. L., & Goldberg, M. E. (1998). Ventral intraparietal area of the macaque: congruent visual and somatic response properties. Journal of Neurophysiology, 79, 126–136.

Farnè, A., & Làdavas, E. (2000). Dynamic size-change of hand peripersonal space following tool use. Neuroreport, 11(8), 1645–1649. 10.1097/00001756-200006050-00010

Farnè, A., Serino, A., & Làdavas, E. (2007). Dynamic size-change of peri-hand space following tool-use: determinants and spatial characteristics revealed through cross-modal extinction. Cortex, 43(3), 436–43. 10.1016/S0010-9452(08)70468-4

Granges, G., Gibson, S. J., Littlejohn, G. O., & Helme, R. D. (1993). Cerebral Event Related Potentials and Heat Pain Thresholds in Fibromyalgia Syndrome. Journal of Musculoskeletal Pain, 1(3-4), 177–184. 10.1300/J094v01n03_17

Granges, G., & Littlejohn, G. (1993). Pressure pain threshold in pain-free subjects, in patients with chronic regional pain syndrome and in patients with fibromyalgia syndrome. Arthritis and Rheumatism, 36(5), 642–646. 10.1002/art.1780360510

Graziano, M. S., & Cooke, D. F. (2006). Parieto-frontal interactions, personal space, and defensive behavior. Neuropsychologia, 44(6), 845–859. 10.1016/j.neuropsychologia.2005.09.009

Graziano, M. S., Hu, X. T., & Gross, C. G. (1997). Visuospatial properties of ventral premotor cortex. Journal of Neurophysiology, 77(5), 2268–2292.

Graziano, M. S., Yap, G. S., & Gross, C. G. (1994). Coding of visual space by premotor neurons. Science, 266, 1054–1057.

Hollins, M., Harper, D., Gallagher, S., Owings, E. W., Lim, P. F., Miller, V., … Maixner, W. (2009). Perceived intensity and unpleasantness of cutaneous and auditory stimuli: an evaluation of the generalized hypervigilance hypothesis. Pain, 141(3), 215–21. 10.1016/j.pain.2008.10.003

Holmes, N. P., Sanabria, D., Calvert, G. A., & Spence, C. (2006). Multisensory interactions follow the hands across the midline: Evidence from a non-spatial visual-tactile congruency task. Brain Research, 1077(1), 108–115. 10.1016/j.brainres.2005.11.010

Kandula, M., Hofman, D., & Dijkerman, H. C. (2014). Visuo-tactile interactions are dependent on the predictive value of the visual stimulus. Neuropsychologia, 70, 358–366. 10.1016/j.neuropsychologia.2014.12.008

Kennett, S., Eimer, M., Spence, C., & Driver, J. (2001). Tactile-visual links in exogenous spatial attention under different postures: convergent evidence from psychophysics and ERPs. J.Cogn Neurosci., 13(0898-929X), 462–478. 10.1162/08989290152001899

Kenward, M. G., & Roger, J. H. (1997). Small Sample Inference for Fixed Effects from Restricted Maximum Likelihood. Biometrics, 53(3), 983–997.

Làdavas, E., di Pellegrino, G., Farnè, a, & Zeloni, G. (1998). Neuropsychological evidence of an integrated visuotactile representation of peripersonal space in humans. Journal of Cognitive Neuroscience, 10(5), 581–589. 10.1162/089892998562988

Lautenschlager, J., Seglias, J., Bruckle, W., & Muller, W. (1991). Comparisons of spontaneous pain and tenderness in patients with primary fibromyalgia. Clinical Rheumatology, 10(2), 168–173. 10.1007/BF02207658

Legrain, V. (2011). Where is my pain? Pain, 152(3), 467–468. 10.1016/j.pain.2010.11.011

Legrain, V., & Torta, D. M. (2015). Cognitive psychology and neuropsychology of nociception and pain. In G. Pickering & S. Gibson (Eds.), Pain, Emotion and Cognition: a Complex Nexus (pp. 3–20). Cham: Springer.

Levitt, H. (1971). Transformed Up-Down Methods in Psychoacoustics. Journal of the Acoustical Society of America, 49, 467–477.

Leys, C., Ley, C., Klein, O., Bernard, P., & Licata, L. (2013). Detecting outliers: Do not use standard deviation around the mean, use absolute deviation around the median. Journal of Experimental Social Psychology, 49(4), 764–766. 10.1016/j.jesp.2013.03.013

Lloyd, D. M., Shore, D. I., Spence, C., & Calvert, G. A. (2003). Multisensory representation of limb position in human premotor cortex. Nature Neuroscience, 6(1), 17–18. 10.1038/nn991

Lorenz, J., & Hypervigilanz, H. (1998). Hyperalgesia or hypervigilance? An evoked potential approach to the study of fibromyalgia syndrome. Zeitschrift Fur Rheumatologie, 57, 19–22. Retrieved from <GO to ISI>://000077892600008

Macaluso, E., Frith, C. D., & Driver, J. (2000). Modulation of human visual cortex by crossmodal spatial attention. Science (New York, N.Y.), 289(5482), 1206–1208. 8750 [pii]

Makin, T. R., Holmes, N. P., Brozzoli, C., Rossetti, Y., & Farnè, A. (2009). Coding of Visual Space during Motor Preparation: Approaching Objects Rapidly Modulate Corticospinal Excitability in Hand-Centered Coordinates. Journal of Neuroscience, 29(38), 11841–11851. 10.1523/JNEUROSCI.2955-09.2009

Makin, T. R., Holmes, N. P., & Zohary, E. (2007). Is That Near My Hand? Multisensory Representation of Peripersonal Space in Human Intraparietal Sulcus. Journal of Neuroscience, 27(4), 731–740. 10.1523/JNEUROSCI.3653-06.2007

Maravita, A., & Iriki, A. (2004). Tools for the body (schema). Trends in Cognitive Sciences, 8(2), 79–86. 10.1016/j.tics.2003.12.008

McDermid, A. J., Rollman, G. B., & McCain, G. a. (1996). Generalized hypervigilance in fibromyalgia: Evidence of perceptual amplification. Pain, 66, 133–144. 10.1016/0304-3959(96)03059-X

Miró, E., Lupiáñez, J., Hita, E., Martínez, M. P., Sánchez, A. I., & Buela-Casal, G. (2011). Attentional deficits in fibromyalgia and its relationships with pain, emotional distress and sleep dysfunction complaints. Psychology & Health, 26(6), 765–780. 10.1080/08870446.2010.493611

Moseley, L. G., Gallace, A., & Spence, C. (2012). Bodily illusions in health and disease: physiological and clinical perspectives and the concept of a cortical “body matrix.” Neuroscience and Biobehavioral Reviews, 36(1), 34–46. 10.1016/j.neubiorev.2011.03.013

Pinheiro, J. C., & Bates, D. M. (2000). Mixed-Effects Models in S and S-PLUS. New York: Springer-Verlag.

Ratcliff, R. (1993). Methods for dealing with reaction time outliers. Psychological Bulletin. 10.1037/0033-2909.114.3.510

Rizzolatti, G., Fadiga, L., Fogassi, L., & Gallese, V. (1997). The space around us. Science, 277, 190–191. 10.1126/science.277.5323.190

Rizzolatti, G., Scandolara, C., Matelli, M., & Gentilucci, M. (1981). Afferent properties of periarcuate neurons in macaque monkeys. I. Somatosensory responses. Behavioural Brain Research, 2, 125–146. 10.1016/0166-4328(81)90052-8

Rollman, G. B., & Lautenbacher, S. (1993). Hypervigilance effects in fibromyalgia: pain experience and pain perception. In H. Vaeroy & H. Merskey (Eds.), Progress in Fibromyalgia and Myofascial Pain (pp. 149–159). Amsterdam: Elsevier.

Sambo, C. F., & Forster, B. (2009). An ERP investigation on visuotactile interactions in peripersonal and extrapersonal space: evidence for the spatial rule. Journal of Cognitive Neuroscience, 21(8), 1550–9. 10.1162/jocn.2009.21109

Schiff, W. (1965). Perception of Impending Collision: a Study of Visually Directed Avoidant Behavior. Psychological Monographs: General and Applied, 79(11), 1 –26. 10.1037/h0093887

Schiff, W., Caviness, J. A., & Gibson, J. J. (1962). Persistent fear responses in rhesus monkeys to the optical stimulus of “looming.” Science, 136(1962), 982–983.

Scudds, R. A., Rollman, G. B., Harth, M., & McCain, G. A. (1987). Pain perception and personality. Journal of Rheumatology, 14, 563–569.

Serino, A., Bassolino, M., Farnè, A., & Làdavas, E. (2007). Extended multisensory space in blind cane users. Psychological Science, 18(7), 642–648. 10.1111/j.1467-9280.2007.01952.x

Spence, C., & Driver, J. (2004). Crossmodal space and crossmodal attention. (C. Spence & J. Driver, Eds.). Oxford: Oxford University Press.

Spence, C., Pavani, F., & Driver, J. (2000). Crossmodal links between vision and touch in covert endogenous spatial attention. Journal of Experimental Psychology, 26(4), 1298–1319.

Spence, C., Pavani, F., & Driver, J. (2004). Spatial constraints on visual-tactile cross-modal distractor congruency effects. Cognitive, Affective & Behavioral Neuroscience, 4(2), 148–169. 10.3758/CABN.4.2.148

Staud, R., Robinson, M. E., & Price, D. D. (2007). Temporal Summation of Second Pain and Its Maintenance Are Useful for Characterizing Widespread Central Sensitization of Fibromyalgia Patients. Journal of Pain, 8(11), 893–901. 10.1016/j.jpain.2007.06.006

Taffou, M., & Viaud-Delmon, I. (2014). Cynophobic Fear Adaptively Extends Peri-Personal Space. Frontiers in Psychiatry, 5(September), 3–9. 10.3389/fpsyt.2014.00122

Taylor-Clarke, M., Kennett, S., & Haggard, P. (2002). Vision modulates somatosensory cortical processing. Curr Biol, 12, 233–236.

Teneggi, C., Canzoneri, E., di Pellegrino, G., & Serino, A. (2013). Social Modulation of Peripersonal Space Boundaries. Current Biology, 23(5), 406–411. 10.1016/j.cub.2013.01.043

Tinbergen. (1951). The study of instinct. Oxford: Clarendon Press.

Tunks, E., Crook, J., Norman, G., & Kalaher, S. (1988). Tender points in fibromyalgia. Pain, 34(1), 11–19. 10.1016/0304-3959(88)90176-5

Vagnoni, E., Lourenco, S. F., & Longo, M. R. (2012). Threat modulates perception of looming visual stimuli. Current Biology, 22(19), R826–R827. 10.1016/j.cub.2012.07.053

Van Damme, S., Van Hulle, L., Spence, C., Devulder, J., Brusselmans, G., & Crombez, G. (2015). Hypervigilance for innocuous tactile stimuli in patients with fibromyalgia: An experimental approach. European Journal of Pain, 19(5), 706–714. 10.1002/ejp.593

Van der Biest, L., Legrain, V., De Paepe, A., & Crombez, G. (2015). Watching what’s coming near increases tactile sensitivity: an experimental investigation. Behavioural Brain Research, 297, 307–314. 10.1016/j.bbr.2015.10.028

Von Korff, M., Ormel, J., Keefe, F. J., & Dworkin, S. F. (1992). Grading the severity of chronic pain. Pain, 50, 133–149.

Wall, P. (1993). The mechanisms of fibromyalgia: a critical essay. In H. Vaeroy & H. Merskey (Eds.), Progress in Fibromyalgia and Myofascial Pain (pp. 53–59). Amsterdam: Elsevier.

West, B. T., Welch, K. B., & Galecki, A. T. (2007). Linear mixed models: A practical guide using statistical software. London: Chapman and Hall/CRC.

Wolfe, F., Clauw, D. J., Fitzcharles, M.-A., Goldenberg, D. L., Katz, R. S., Mease, P., … Yunus, M. B. (2010). The American College of Rheumatology Preliminary Diagnostic Criteria for Fibromyalgia and Measurement of Symptom Severity. Arthritis Care & Research, 62(5), 600–610. 10.1002/acr.20140

